# Humeral elevation workspace during daily life of adults with spinal cord injury who use a manual wheelchair compared to age and sex matched able-bodied controls

**DOI:** 10.1101/2020.07.31.20138305

**Authors:** Brianna M. Goodwin, Stephen M. Cain, Meegan G. Van Straaten, Emma Fortune, Melissa M. B. Morrow

## Abstract

Shoulder pain and pathology are extremely common for individuals with spinal cord injuries (SCI) who use manual wheelchairs (MWC). Although risky humeral kinematics have been measured during wheelchair-based activities performed in the lab, little is known about arm kinematics in the free-living environment. The purpose of this study was to measure the humeral elevation workspace throughout a typical day for individuals with SCI who use a MWC and matched able-bodied controls. Thirty-four individuals with SCI who use a MWC (42.7±12.7 years of age, 28 males/6 females, C6-L1) and 34 age- and sex-matched controls were enrolled. Participants wore three inertial measurement units (IMU) on their upper arms and torso for one to two days. Humeral elevation angles were estimated and the percentage of time individuals spent in five elevation bins (0-30°, 30-60°, 60-90°, 90-120°, and 120-180°) were calculated. For both arms, the SCI cohort spent a significantly lower percentage of the day in 0-30° of humeral elevation (Dominant: SCI= 15.7±12.6%, Control= 32.1±15.6%, p<0.0001; Non-Dominant: SCI= 21.9±17.8%, Control= 34.3±15.5%, p=0.001) and a significantly higher percentage of time in elevations associated with tendon compression (30-60° of humeral elevation, Dominant: SCI= 62.8±14.4%, Control= 49.9.1±13.0%, p<0.0001; Non-Dominant: SCI= 58.8±14.9%, Control= 48.3±13.6%, p=0.003) than controls. The increased percentage of time individuals with SCI spent in elevations associated with tendon compression may contribute to increased shoulder pathology. Characterizing the humeral elevation workspace utilized throughout a typical day may help in understanding the increased prevalence of shoulder pain and pathology in individuals with SCI who use MWCs.

## Introduction

Shoulder pain is the most common site of musculoskeletal pain in adults with spinal cord injuries (SCI) who use manual wheelchairs (MWC) and its existence can significantly limit a person’s functional abilities (1). Shoulder pain is reported in 37-70% of individuals with SCI who use a MWC (2-7). This differs vastly from the 2.9% of the general able-bodied population who experience shoulder pain (8). Although shoulder pain can develop any time after SCI, it is most commonly developed within the first five years (9) and often lasts longer than one year (3). Of the MWC users who experience pain, up to 93% have pathological signs on MRI (10), most commonly in the supraspinatus tendon (11).

In general, non-traumatic supraspinatus tendon tears in the shoulder have been thought to be caused by a combination of intrinsic and extrinsic factors (12). However, these effects can be exacerbated by overuse (13). One extrinsic factor is the narrowing of the subacromial space which causes compression of the supraspinatus tendons under the coracoacomial arch, and is hypothesized to lead to increased tendon pathology and pain (14, 15). Individualized musculoskeletal models utilizing MRI have estimated the risk of supraspinatus tendon compression through various humeral planes and elevations (16). The magnitude of glenohumeral elevation was the greatest kinematic predictor of tendon compression risk, followed by the specific plane of elevation. Results showed the supraspinatus tendon had the greatest risk of compression at humerothoracic elevations angles between 30-60° (15). Biplane fluoroscopic imaging of the shoulder joint during dynamic motion has shown similar results and demonstrated that at higher humeral elevations, as the humeral head rotates posteriorly, the supraspinatus tendon may no longer be under the coracoacromial arch, therefore, not at risk of compression (17). Understanding where tendon compression risk occurs can provide insights when interpreting the humeral elevation workspace of activities of daily living.

MWC propulsion, transfers, and other wheelchair-based activities of daily living have been investigated in laboratory environments to characterize the upper extremity kinematics that pose a risk for shoulder tendon compression (18-20). Although in-laboratory data provide accurate quantifications of how MWC users utilize their arms to complete specific activities, it is unable to quantify the exposure to potentially risky postures in daily-living. To understand the daily exposure to risky shoulder motion, inertial measurement units (IMUs) can be used to measure the angular velocity and acceleration of body segments throughout an entire day in environments of daily living. IMU-based methods for quantifying shoulder movement show good agreement with position-based motion capture and have been used to quantify shoulder elevation angles; however a limited number of studies have applied these methods to free-living full-day collections (21-26). To the best of our knowledge no study has utilized these methods to understand the humeral elevation workspace of MWC users throughout an entire day.

The purpose of this study was to use IMUs to measure the humeral elevation workspace throughout a typical day for individuals with SCI who use a MWC and compare it to matched able-bodied controls. Comparison to controls allows for understanding of how humeral elevation exposure during daily life differs when the option to use the lower extremities for weight bearing and mobility is removed. This study also aimed to understand the effects of years of MWC use, pain, sex, and level of SCI on the humeral elevation workspace. Due to the increased prevalence of shoulder pain and pathology in MWC users compared to able-bodied controls (11) and the potential role that humeral elevation has on shoulder tendon compression (15), we hypothesized that MWC users would utilize a different humeral elevation workspace than able-bodied adults. Specifically, we hypothesized individuals with SCI would spend a higher percentage of time at elevation angles associated with tendon compression risk. Understanding the humeral elevation workspace of individuals with SCI may contribute to understanding why increased levels of shoulder pain and pathology occur for this population.

## Methods

### Participant Enrollment

This study was approved by the Mayo Clinic Institutional Review Board. Individuals with an SCI who used a MWC as their main mode of mobility were recruited through querying medical records and care providers of local clinics. Sex- and age- (±2.5 years) matched able-bodied controls were recruited through email distribution lists and classified ads. Participants for both cohorts were considered for inclusion in the study if they were between 18-70 years of age and had functional range of motion at both shoulders. Functional range of motion was defined as active humeral thoracic flexion, abduction of at least 150° and the ability of the participant to touch the opposite shoulder, the back of his/her neck and his/her low back. Prior to accrual to the study a licensed physical therapist (MVS) performed a screening physical exam to confirm inclusion and exclusion criteria. Participants with SCI were excluded if they self-reported previous diagnoses of complete supraspinatus tendon tears bilaterally. Controls were excluded with a known unilateral or bilateral complete supraspinatus tendon tear(s). Additionally, participants in both cohorts were excluded if there were conditions/factors which might have hindered protocol adherence and controls were also excluded if they had any musculoskeletal or neurological disorder which might have impacted shoulder health or changed the individual’s ability to walk independently.

### Questionnaires and IMU Instrumentation

Upon enrollment, participants attended an in-lab visit. A licensed physical therapist screened participants for eligibility and informed consent was obtained. Participants self-reported their hand dominance and were asked if they had pain on either or both shoulders. All participants completed the Disabilities of the Arm, Shoulder, and Hand (DASH) questionnaire (27) for both right and left arms. The DASH encompasses 30 questions which ask individuals to rate their difficulty, pain, and satisfaction when accomplishing specific tasks on a 5 point scale. Scores range from 0-100, with 0 indicating no difficulty and 100 indicating the most difficulty, pain, and dissatisfaction. The DASH has been shown to be reliable and to have high validity (28). Additionally, the SCI cohort filled out the Wheelchair User Shoulder Pain Index (WUSPI) for both the right and left shoulders. To complete the WUSPI, participants were asked to rate their shoulder pain when completing 15 tasks on a visual analog scale between “no pain” and “worst pain ever experienced” (29). Possible scores ranged from 0 (no pain) and 150 (worst pain ever experienced in all categories). The WUSPI is valid and reliable for this population (30). Although we acknowledge that the DASH and WUSPI were designed to be filled out once, as part of a larger study, both surveys were filled out for both arms to evaluate pain and function as it related to each arm

Participants were given three wireless IMUs (Emerald or Opal, APDM, Inc., Portland, OR). Each IMU contained a 3-axis accelerometer (±200 g), 3-axis gyroscope (±2000°/s), and 3-axis magnetometer (±8 Guass). The three IMUs remained synchronized via a proprietary wireless protocol, recorded data at 128 Hz and saved the data to internal storage. In order to maximize the consistency of IMU placement and functional calibration movements across participants, written handouts, video guides and in-person instruction were provided. Participants were instructed to wear one IMU on each lateral upper arm and one on the anterior of the torso; IMUs were secured on the body with elastic and Velcro straps. Each IMU was labeled with the wear location (left arm, right arm, or torso) and an arrow indicating the proper mounting orientation. Participants were instructed to wear the sensors during the entire length of two typical days, excluding bathing and swimming, and take them off before going to bed. Both cohorts were asked to perform their regular daily routines; participants in the control cohort did not use MWCs. Upon donning the sensors for a day, participants performed a set of functional calibration postures (Fig 1, Appendix A). Due to the collection of multiple days of data, participants were responsible for charging the IMUs overnight using a provided charging station. After the data collection, participants returned the sensors with a pre-paid mailer or in person to the study staff.

**Fig 1:**
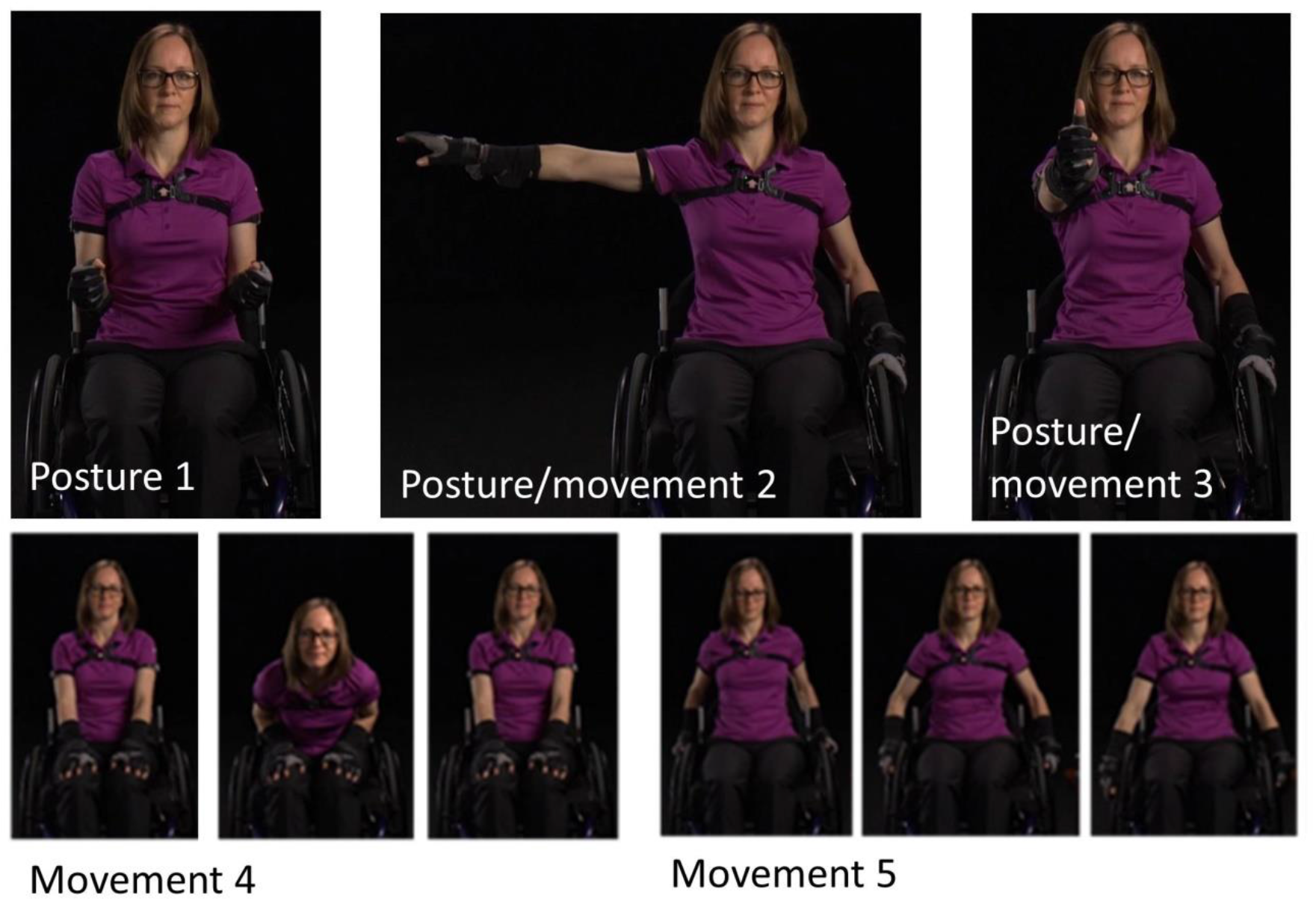
Functional calibration used to align IMU’s with the body. Postures included static upright neutral posture with upper arms resting against the thorax (posture 1), static and dynamic arm t-pose/movement (shoulder abduction = 90°, posture 2), static and dynamic flexion pose/movement (shoulder flexion = 90°, posture 3), dynamic flexion and extension of the torso (movement 4), and simulated wheelchair use or walking (movement 5). Postures 2 and 3 were completed for both the right and left arms separately. (Note: The individual pictured is a co-author).

### Data Processing

Data were downloaded through Motion Studio (APDM, Inc., Portland, OR) and outputs included estimates of the orientations of each IMU relative to an inertial frame (Fig 2). The orientation estimates were derived from the combined acceleration and angular velocity data, rather than only the acceleration data. While researchers have used IMU-measured acceleration only to estimate arm orientation (31, 32), there are known limitations to this approach (33), namely, the challenge of separating the measured acceleration into gravitational and body caused components. Algorithms that use both measured acceleration and angular velocity to estimate IMU orientation or attitude (orientation relative to gravity) are well understood and are critical in strapdown inertial navigation (34). These algorithms integrate the angular velocity signal to estimate orientation during periods with high dynamics (significant body acceleration) and use the acceleration signal to update or correct the orientation during periods with low dynamics (measured acceleration close to the acceleration of gravity). Further, these algorithms take different forms and have been proven to be highly accurate for estimating attitude (35-38). Additionally, the orientation estimates were calculated without magnetometer data due to the unknown and likely non-uniform magnetic fields present throughout field data collections. While the orientation algorithm used by APDM is proprietary, sensor fusion methods (e.g. Kalman filters) used to estimate IMU orientation from raw sensor data are well understood and well documented in the literature (21, 34, 35). Custom MATLAB (Mathworks, Natick, MA) code was written to calculate orientations of anatomical axes relative to IMU-fixed reference frames using data collecting during each participant’s functional calibration postures and movements (Fig 1; Appendix A). Orientation of a given body segment (upper arm or thorax) in an inertial (world) reference frame was then estimated using the orientation of the IMU and the orientation of the anatomical axes relative to the IMU-fixed reference frame (Appendix B). Humeral elevation and thorax deviation angles were defined as the angle between the long axis of the body segment (defined from the function calibration) and vertical; these angles are only dependent on the estimated direction of gravity relative to the body segment and, therefore, are drift-free metrics for quantifying body segment motions. The calculated humeral elevation angles range between 0-180°, with 0° indicating the arm was down and perfectly aligned with gravity and 180° indicating the arm was raised overhead and aligned with gravity. These methods have previously been validated in unpublished data where five individuals with SCI performed 10 reaching tasks. The absolute error and percent of error when compared to the gold standard (electromagnetic system) were -0.06±1.12° and -1.44±1.28%, respectively, for the range of motion. The absolute error and percent of error for the maximum elevation achieved during each reach were 2.59±2.47° and 2.04±2.47%, respectively.

**Fig 2:**
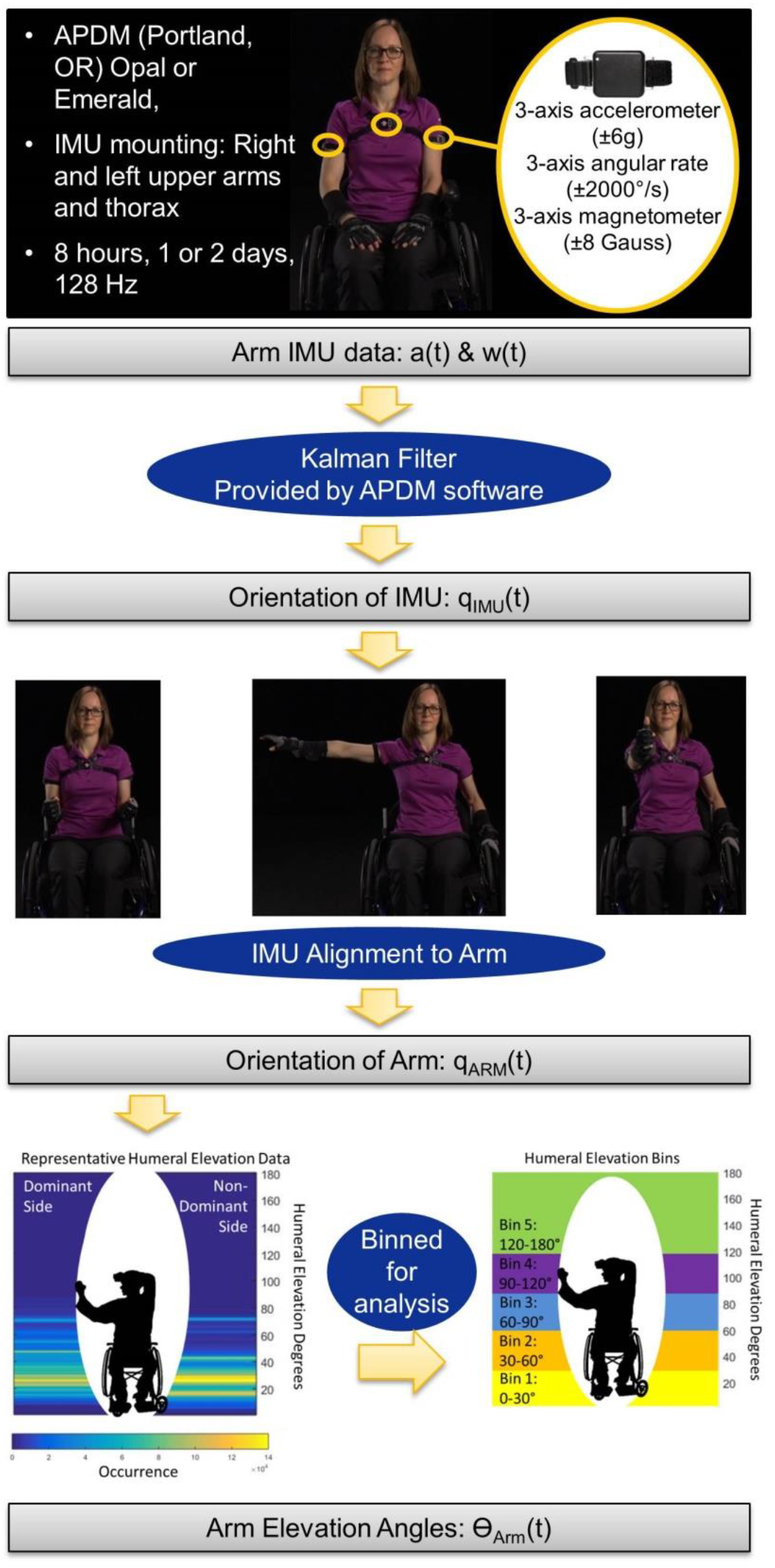
Data processing workflow. This included data collected, IMU orientation, IMU alignment to arm through calibration postures, representative data, and humeral elevation bins. The percentage of time spent in each bin was calculated and used for analysis. (Note: The individual pictured is a co-author).

It is important to note that humerothoracic elevation angles and elevation planes relative to the thorax were not calculated as these calculations require accounting for relative drift between the orientation estimates of arm and torso IMUs. While the attitude estimates are accurate and do not drift, the yaw or heading estimates do drift, making accurate calculation of humerothoraic angles over long periods of time difficult (24). This difficulty is best illustrated by the fact that studies that use IMUs to quantify shoulder motion during long periods in the real world either do not calculate shoulder angles(31, 39-41) or acknowledge the limitations of the methodology (24). Other work (25, 42) claims to accurately calculate shoulder angle of elevation but not plane of elevation; however, shoulder angle of elevation cannot be calculated accurately without the plane of elevation (43). Correcting the drift between sensors about vertical is an active research area and requires a joint specific approach (44, 45). Therefore, in our analysis, data in which the thorax deviation angle was more than 30° were eliminated in order to allow humeral elevation angles to be interpreted similarly to humerothoracic elevation angles; 30° was selected based on a sensitivity analysis performed during a prior study.

The percentage of daily wear time each participant spent in five humeral elevation bins were calculated (0-30°, 30-60°, 60-90°, 90-120°, and 120-180°). The bin sizes were chosen as a means to combine three theories: 1) a painful arc of motion occurs between 60-120° of arm abduction (46), 2) Rapid Upper Limb Assessment (RULA) which bins risky arm postures between 0-20°, 20-45°, 45-90°, and >90° (47), and 3) the subacromial risk area of 30-60° (15).

Periods of non-wear were determined using methods from Lugade and colleagues (2014) and were excluded from data analysis. Data were also excluded from analysis if the functional calibration postures were not completed properly or if at least eight hours of data were not collected after the elimination of non-wear time. Data were included if one or two complete days were collected; if two days were included all data were combined before the calculation of the percent of time in humeral elevation bins.

### Statistical Analysis

Multivariate analyses of variance (MANOVA) were used to test for the main effects of cohort, sex, age, pain (measured by the WUSPI and DASH), level of injury, and years of MWC use on both dominant and non-dominant side humeral elevation bins (α=0.05). To test for level of injury the SCI cohort was divided into three groups based on their injury level; cervical (C6-C8), high/mid thoracic (T1-T8), and low thoracic/lumbar (T9-L1). Within groups, analysis of variance (ANOVA) was used to test for the main effect of humeral elevation bin for both arms. When significant main effects were observed, post hoc paired t-tests were performed. A Bonferroni correction factor was used to adjust the alpha level from 0.05 to 0.01 due to the calculation of time in five bins.

## Results

Thirty-four participants with SCI who used a MWC, and 34 age (±2.5 years) and sex matched, able-bodied adults were enrolled (Table 1). There were no statistical differences between the cohort’s self-reported weight, height, and dominant hand.

**Table 1:** Participant Demographics.

|  | **SCI** | **Control** | **P-value** |
| --- | --- | --- | --- |
| **Age** | 42.7 +/- 12.7<br>(22.6 - 63.3) | 42.6 +/- 12.5<br>(24.3 - 61.0) | 0.776 |
| **Sex** | 28 males/6<br>females | 28 males/6 females | - |
| **Self-reported weight (kg)** | 80.7 +/- 17.2<br>(54.0 - 145.1) | 81.6 +/- 17.5<br>(56.7 - 149.7) | 0.6822 |
| **Self-reported height (cm)** | 177.4 +/- 7.6<br>(160.0 -<br>195.6) | 178.4 +/- 9.5<br>(160.0 - 205.7) | 0.417 |
| **Injury Level** |  |  |  |
| Cervical (C6-C8) | 7 | - | - |
| High/mid thoracic (T1-T8) | 16 |  |  |
| Low thoracic/lumbar (T9-L1) | 11 |  |  |
| **Years of manual wheelchair**<br>**use (years)** | 11.5 +/- 10.7<br>(0.5 - 36.0) | - | - |
| **Dominant arm** | 27 right/7 left | 32 right/2 left | 0.058 |
| **DASH (dominant arm)** | 15.2 +/- 17.5<br>(0 - 71.7) | 1.3 +/- 2.9<br>(0 - 15) | <0.0001 |
| **DASH (non-dominant arm)** | 13.6 +/- 14.2<br>(0 - 51.7) | 1.1 +/- 3.3<br>(0 - 15) | <0.0001 |
| **WUSPI (dominant arm)** | 12.7 +/- 20.4<br>(0 - 71.2) | - | - |
| **WUSPI (non-dominant arm)** | 10.8 +/- 16.2<br>(0 - 71.6) | - | - |
| **Presence of pain** | 26 | 9 | <0.0001 |
| **Number of participants** | 34 | 34 | - |

### Excluded Data

One control was excluded from the study due to complete supraspinatus tear; however no SCI participants were excluded due to the previous diagnosis of a complete supraspinatus tear. Seven pairs of data were excluded from the analysis due to exclusion criteria (Fig 3). Data were collected for an average (SD) of 11.4(2.1) and 11.9(1.3) hours for the SCI and control cohorts, respectively. Additionally, on average 18.3(14.0) and 28.0(10.3) percent of the day was excluded because the trunk was at or over 30° for the SCI and control cohorts, respectively.

**Fig 3:**
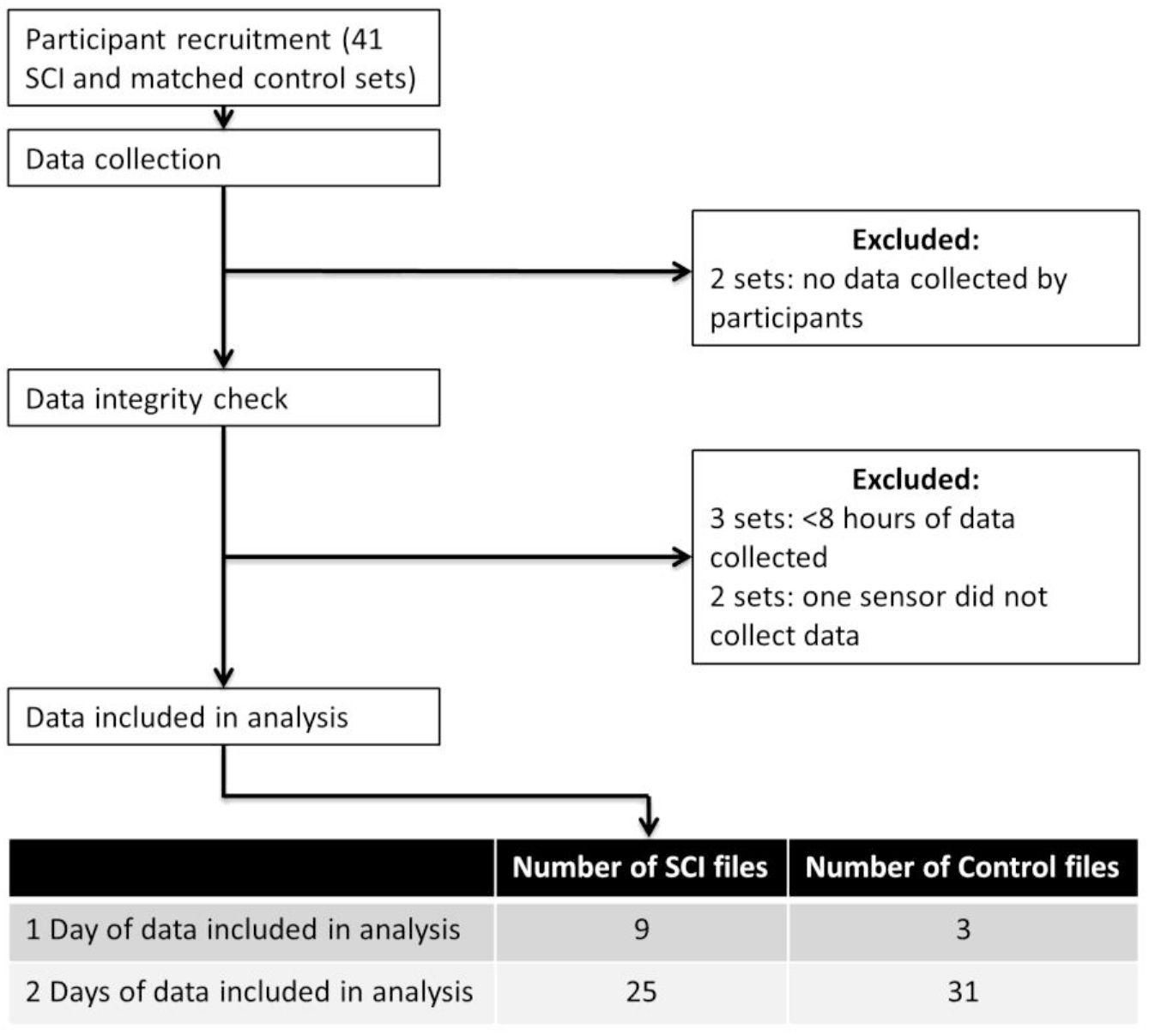
Data exclusion processes. Data was excluded if either SCI or control did not collect data, a minimum of 8 hours of data were not collected, or one sensor malfunctioned. Data was included in analysis if one or two days of data was collected.

### Humeral Elevation Workspace

There was a main effect of cohort across humeral elevation bins on both dominant and non-dominant sides (p<0.0001). Additionally, there was a main effect of humeral elevation bin for both cohorts and arms (dominant: p<0.0001, non-dominant: p=0.005, Fig 4). Individuals with SCI spent significantly more time in 30-60° of humeral elevation than all other elevations bins on both their dominant and non-dominant sides (p<0.001, Table 2). The SCI cohort spent 63% and 59% of their daily wear time (approximately 7 hours per day) at these elevations on their dominant and non-dominant sides, respectively. The controls also spent the greatest amount of daily wear time in this elevation bin at 50% and 48% on their dominant and non-dominant arm respectively, which was significantly lower than the SCI cohort for both arms (dominant: p<.0001, non-dominant: p=0.003, Table 2).

**Fig 4:**
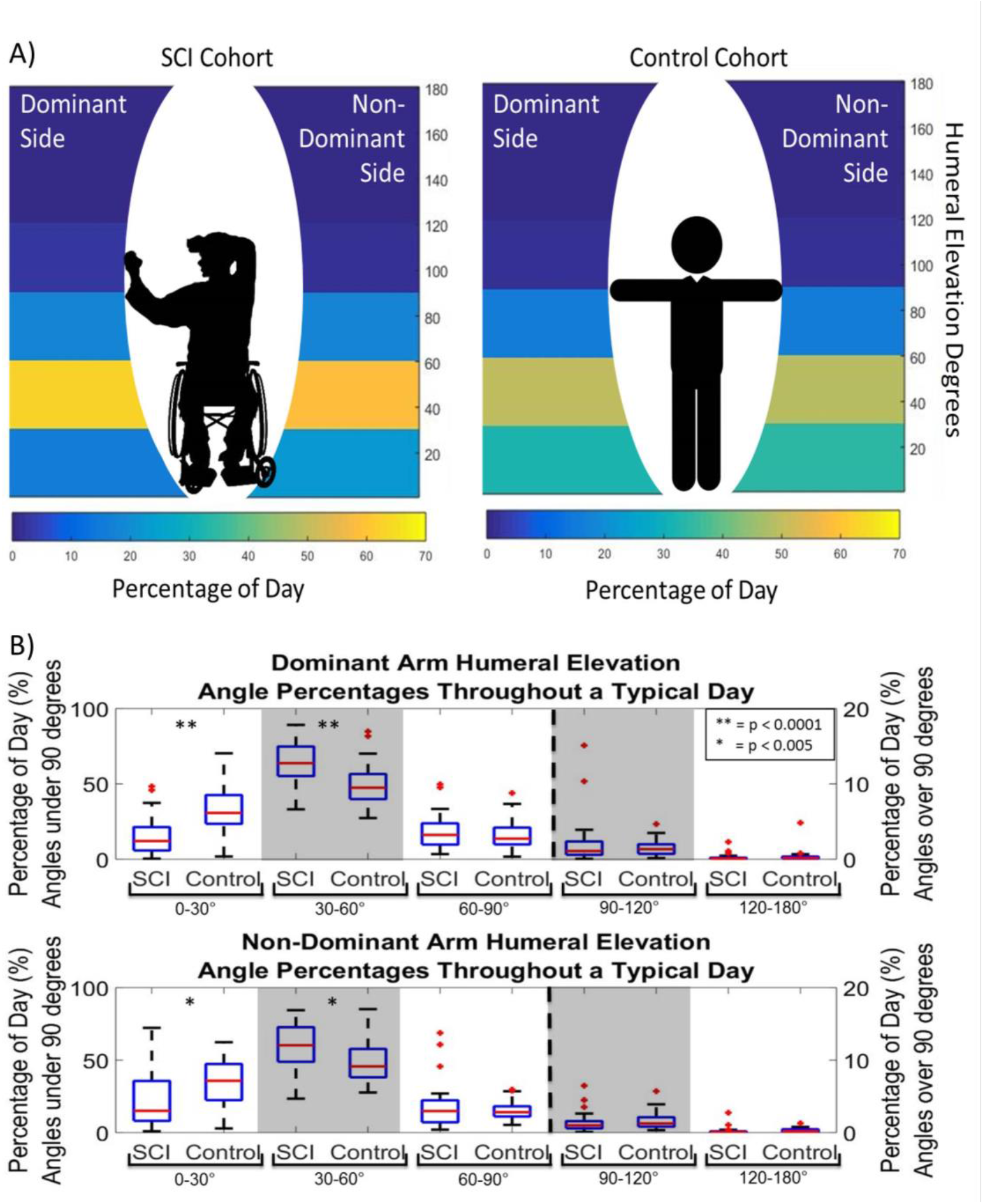
The percentage of time in each humeral elevation. A) The average percentage of time throughout a typical day individuals in the SCI and control cohorts spent in 0-30°, 30-60°, 60-90°, 90-120°, and 120-180° of humeral elevation for their dominant and non-dominant sides. B) Percentage of time throughout a typical day individuals in the SCI and control cohorts spent in 0-30°, 30-60°, 60-90°, 90-120°, and 120-180° of humeral elevation for their dominant arm (top) and their non-dominant arm (bottom). For each boxplot the central line (red) represents the median, the edges of the box are the 25 ^th^ and 75 ^th^ percentiles, and the error bars extend the most extreme data points not considered outliers and, the outliers are denoted by red +. ** indicates p < 0.0001 and * indicates p < 0.005.

**Table 2:** The average (SD) percentage of the day individuals with SCI and matched able-bodied controls spent in five humeral elevation bins throughout one or two days.

| **Bin** | **SCI Dominant Hand Percentage (%)** | **Control Dominant Hand Percentage (%)** | **P-Value** | **SCI Non-Dominant Arm Percentage (%)** | **Control Non-Dominant Arm Percentage (%)** | **P-Value** |
| --- | --- | --- | --- | --- | --- | --- |
| **0-30°** | 15.7 (12.6) | 32.1 (15.6) | <0.0001 | 21.9 (17.8) | 34.3 (15.5) | 0.001 |
| **30-60°** | 62.8 (14.4) | 49.9 (13.0) | <0.0001 | 58.8 (14.9) | 48.3 (13.6) | 0.003 |
| **60-90°** | 18.4 (11.0) | 16.2 (9.6) | 0.410 | 17.7 (14.8) | 15.6 (6.2) | 0.430 |
| **90-120°** | 2.8 (5.3) | 1.4 (1.0) | 0.145 | 1.4 (1.4) | 1.6 (1.2) | 0.589 |
| **120° - 180°** | 0.2 (0.4) | 0.4 (0.8) | 0.320 | 0.2 (0.5) | 0.3 (0.3) | 0.430 |

For the SCI cohort, the second largest percentage of time was spent in 60-90° of humeral elevation (approximately 20% of their day for both arms). Controls spent their second largest percentage of time in 0-30° of elevation for both arms, which was significantly higher than the amount of time the SCI cohort spent in this elevation bin (p<0.001). Individuals with SCI spent comparable amounts of time in 0-30° and 60-90° of elevation, while controls spent significantly more time in 0-30° than 60-90° of humeral elevation on the dominant (p<0.001) and non-dominant (p<0.0001) sides.

On average, participants in both cohorts spent less than 3% of their day (<25 minutes) in elevations over 90° for both arms. There were no significant differences between cohorts for the 60-90° 90-120°and >120° humeral elevation bins or between dominant and non-dominant arms for each cohort and each elevation bin.

### Pain

Pain, measured by the DASH and WUSPI, did not have a significant effect on the percentage of time an individual spent in any humeral elevation bins for both dominant and non-dominant arms (p>0.01).

### Sex, Age, Injury level, and Years of MWC Use

There were no main effects of sex (Table 3), age (Table 4), injury level (Table 3), or years of MWC Use (Table 4) on either arm (p>0.01).

**Table 3:**
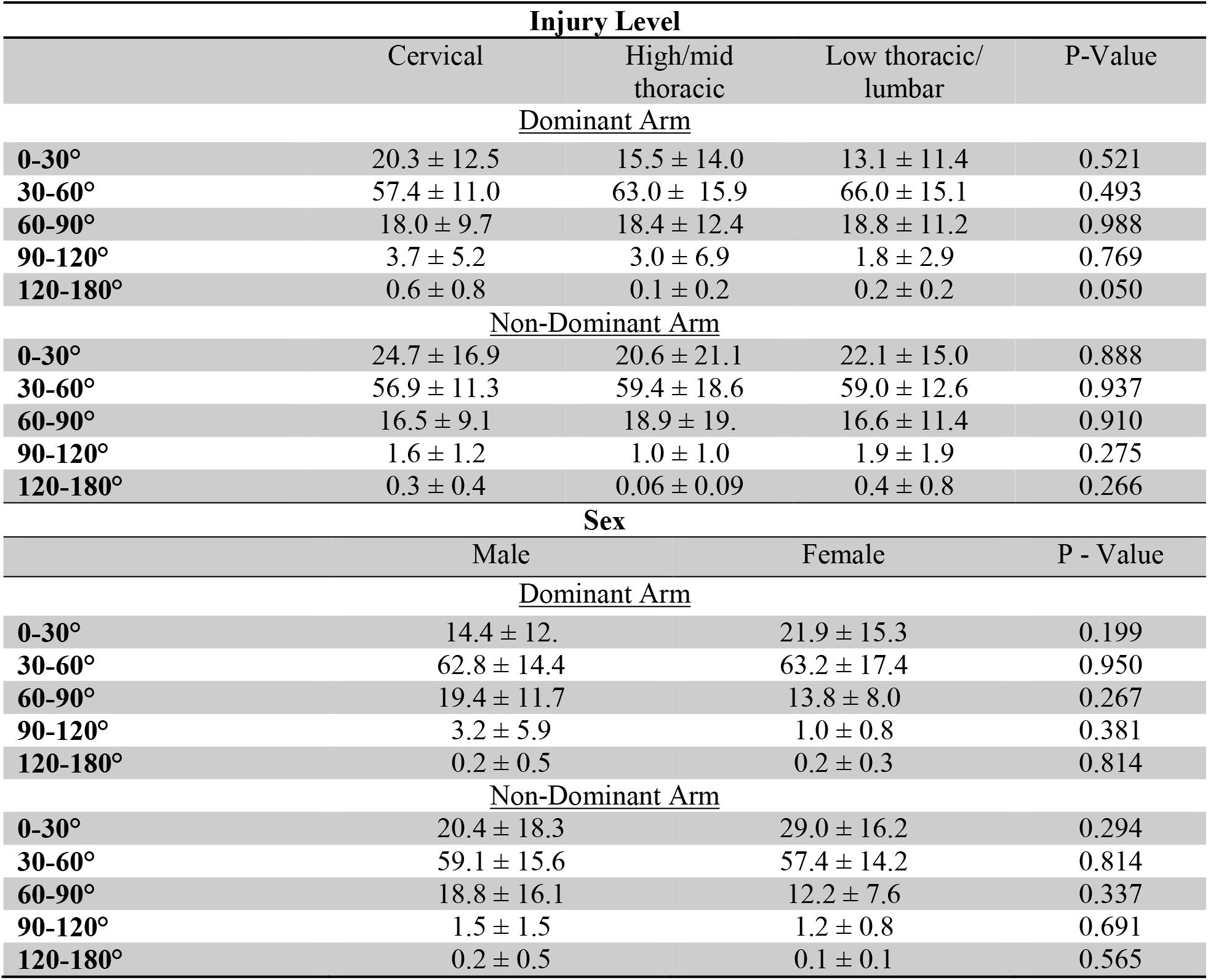
The percentage of time individuals spent in humeral elevation bins based on their injury level and sex.

| Injury Level |  |  |  |  |
| --- | --- | --- | --- | --- |
|  | Cervical | High/mid thoracic | Low thoracic/ lumbar | P-Value |
| Dominant Arm |  |  |  |  |
| 0-30° | 20.3 ± 12.5 | 15.5 ± 14.0 | 13.1 ± 11.4 | 0.521 |
| 30-60° | 57.4 ± 11.0 | 63.0 ± 15.9 | 66.0 ± 15.1 | 0.493 |
| 60-90° | 18.0 ± 9.7 | 18.4 ± 12.4 | 18.8 ± 11.2 | 0.988 |
| 90-120° | 3.7 ± 5.2 | 3.0 ± 6.9 | 1.8 ± 2.9 | 0.769 |
| 120-180° | 0.6 ± 0.8 | 0.1 ± 0.2 | 0.2 ± 0.2 | 0.050 |
| Non-Dominant Arm |  |  |  |  |
| 0-30° | 24.7 ± 16.9 | 20.6 ± 21.1 | 22.1 ± 15.0 | 0.888 |
| 30-60° | 56.9 ± 11.3 | 59.4 ± 18.6 | 59.0 ± 12.6 | 0.937 |
| 60-90° | 16.5 ± 9.1 | 18.9 ± 19. | 16.6 ± 11.4 | 0.910 |
| 90-120° | 1.6 ± 1.2 | 1.0 ± 1.0 | 1.9 ± 1.9 | 0.275 |
| 120-180° | 0.3 ± 0.4 | 0.06 ± 0.09 | 0.4 ± 0.8 | 0.266 |
| Sex |  |  |  |  |
|  | Male | Female | P - Value |  |
| Dominant Arm |  |  |  |  |
| 0-30° | 14.4 ± 12. | 21.9 ± 15.3 | 0.199 |  |
| 30-60° | 62.8 ± 14.4 | 63.2 ± 17.4 | 0.950 |  |
| 60-90° | 19.4 ± 11.7 | 13.8 ± 8.0 | 0.267 |  |
| 90-120° | 3.2 ± 5.9 | 1.0 ± 0.8 | 0.381 |  |
| 120-180° | 0.2 ± 0.5 | 0.2 ± 0.3 | 0.814 |  |
| Non-Dominant Arm |  |  |  |  |
| 0-30° | 20.4 ± 18.3 | 29.0 ± 16.2 | 0.294 |  |
| 30-60° | 59.1 ± 15.6 | 57.4 ± 14.2 | 0.814 |  |
| 60-90° | 18.8 ± 16.1 | 12.2 ± 7.6 | 0.337 |  |
| 90-120° | 1.5 ± 1.5 | 1.2 ± 0.8 | 0.691 |  |
| 120-180° | 0.2 ± 0.5 | 0.1 ± 0.1 | 0.565 |  |

**Table 4:** The linear regression values for the percentage of time individuals spent in humeral elevation bins based on age and years of MWC use.

|  | Age |  | Years of MWC use |  |
| --- | --- | --- | --- | --- |
|  | R<sup>2</sup> | P-Value | R<sup>2</sup> | P-Value |
| <u>Dominant Arm</u> |  |  |  |  |
| **0-30°** | 0.104 | 0.06 | 0.009 | 0.60 |
| **30-60°** | 0.037 | 0.28 | 0.000 | 0.97 |
| **60-90°** | 0.013 | 0.41 | 0.008 | 0.62 |
| **90-120°** | 0.000 | 0.94 | 0.002 | 0.82 |
| **120-180°** | 0.002 | 0.79 | 0.047 | 0.22 |
| <u>Non-Dominant Arm</u> |  |  |  |  |
| **0-30°** | 0.027 | 0.35 | 0.001 | 0.84 |
| **30-60°** | 0.002 | 0.81 | 0.009 | 0.58 |
| **60-90°** | 0.025 | 0.37 | 0.007 | 0.65 |
| **90-120°** | 0.000 | 0.88 | 0.054 | 0.19 |
| **120-180°** | 0.049 | 0.21 | 0.023 | 0.39 |

## Discussion

This study aimed to understand the humeral elevation workspace utilized throughout a typical day by individuals with SCI who use a MWC. These results were compared to controls to better understand factors which may contributed to a higher rate of both pain and tendon pathology associated with years of MWC use (11). Both individuals with SCI and controls spent the majority of their day (~80%) in elevation angles between 0 and 60°. However, individuals with SCI spent significantly more time in humeral elevations associated with supraspinatus tendon compression (30-60°) than controls. There was no evidence of the effect of injury level, years of MWC use, age, or sex on the humeral elevation workspace individuals with SCI spend time in.

With the growing capabilities of wearable technology, many SCI-specific algorithms have been created and validated to accompany and enhance data captured in a lab setting (49). Many of the studies using wearable technology to understand movement of MWC users have focused specifically on wheelchair propulsion and use (50, 51), with less focus on understanding humeral elevation angles or overuse of the arms of MWC users. The data presented in the current study supplements data collected in a laboratory setting and other free-living MWC use metrics by providing lengths of exposure to risky postures in the free living environment.

Recently it has been suggested that compression of the supraspinatus tendon occurs at low elevation angles. Giphart, van der Meijden (17) suggested that subacromial impingement syndrome occurs below 70° of humeral elevation and the minimum distance between the footprint of the supraspinatus tendon and greater tuberosity occurred between 36° and 65° of humeral elevation. Additionally, using individualized bone models (from MRI) and group averaged kinematics, Lawrence, Schlangen (15) used musculoskeletal simulation models to suggest the minimum distance between the coracoacomial arch and humerus occurred at 42° of humerothoracic elevation. Our results show that individuals with SCI who use a MWC spent significantly more time than controls in a similar range of humeral elevations (30-60°). This difference could be in part due to differences in the arm elevation workspace during mobility. During MWC propulsion the humeral elevation required is between 25 and 55° at a self-selected speed (52, 53); however during walking the humeral elevation angles required are much lower (54). The difference in humeral elevation during mobility likely is not the only contributor to this increase, as MWC users move about 3 km less than able-bodied individuals and only spend a small amount of their day actually propelling themselves; estimates range from 16 to 54 minutes per day (50, 55). Another contributing factor to this discrepancy may be wheelchair setup; for example MWC users may not place their arms in a neutral resting position (0-30°) due to the location of their arm rest. In addition to the humeral elevation workspace differing during propulsion for MWC users and walking for able-bodied individuals, the loading of the shoulder is also different during these two tasks and likely contributes to the increase in pathology in MWC users. Further, additional data collections and analyses are needed to fully understand the clinical implications of the differences in humeral elevation between the wheelchair users and able-bodied control group. Future work will determine more details about these differences such as the activity levels and/or participant activities while in 30-60° of humeral elevation. These future analyses can guide whether a reduction of time spent in these elevation angles should be considered as a possible intervention to prevent or treat shoulder dysfunction.

Capturing a holistic view of an individual’s exposure to potentially risky humeral elevation is dependent on many factors including occupation and activities performed throughout a day. A study looking at 10 able-bodied elderly adults using only accelerometery data found that less than 4% of an individual’s day was spent in elevations above 90°, with the average elevation angle occurring at 40° (42). These results are very similar to the data presented in the current study for both cohorts; about 3% of the day was spent in elevations over 90°. Previous reports have suggested that extended periods of time in overhead motion may be the cause of increased shoulder pain. Our results paired with the most recent modeling and imaging data may suggest that injury to the supraspinatus tendon due to tendon compression of the SCI cohort occurs due to increased time between 30-60° of humeral elevations. Further, pain in higher elevation angles may be caused by other mechanisms (15). Continuing to map this workspace for individuals with SCI who use a MWC while they perform specific tasks (i.e., propulsion or transfers) may help us to further understand daily risk exposures and the contribution of specific tasks. Although out of the scope of this study, activity detection is an active area of research for our group (55).

Multiple challenges exist when using unsupervised real-world IMU data. First, accounting for and correcting the drift of IMU-based body segment orientation estimates is a common challenge in understanding the relative orientation of body segments (i.e. joint angles), especially for extended data collections (see excellent discussion in (24)). The current algorithms utilized in this study do not take the plane of motion into account; 30° of humeral elevation in front of the body, to the side, or behind would all be interpreted as 30° of humeral elevation and are indistinguishable. While we could have used the orientation estimates to calculate humerothoracic angle of elevation and plane of elevation, we know that those calculations would contain errors from the heading/yaw drift. Heading drift directly affects the plane of elevation and accurately calculating the plane of elevation is critical to accurately calculating humerothoracic angle of elevation. Therefore, the data presented here only used the angle of the humerus relative to vertical (humeral elevation angle) and not the trunk (humerothoracic angle). This was compensated for by eliminating humeral elevation time points where the trunk angle was at or over 30° of tilt; participants may have been leaning over or lying down. On average about 10% more data was eliminated from the control data sets than the SCI data sites, indicating the controls had more variability and movement of their trunk than the SCI cohort. Even with these limitations, the methods used in this study to estimate sensor orientation and humeral elevation are more accurate than other methods using only acceleration data, especially during movements with high dynamics (35-38).

There are limitations with the data presented in this study to consider. Previous studies have found that up to four days of data collection are needed to represent propulsion trends consistent throughout a MWC user’s daily life (56). Only one or two days of data were collected for participants in this study due to participant availability and adherence to the protocol. We attempted to compensate for this by asking participants to wear the sensors on ‘typical days.’ The calibration protocol used in this study enabled us to determine humeral elevations without an in-lab calibration. As participants performed the calibration protocol unsupervised, it’s possible that there could be errors induced by incorrect neutral and 90° calibration postures. The data presented here were binned into 30° ranges below 120° of humeral elevation; however, creating bins with different boundaries may affect the results. Appendix C shows the average percent of time in 10° bins. Additionally, there are other factors beyond humeral elevation that contribute to shoulder injury in the SCI population including increased load on the shoulder due to MWC propulsion, body transfers, and repetitive motion. Loading of the shoulder although not measured in this study, has an important role in the increased pathology and pain for MWC users.

## Conclusions

This study aimed to understand the humeral elevation workspace throughout a typical day of individuals with SCI who use a MWC and compare it to the workspace of age- and sex-matched controls. Our data suggest that individuals with SCI who use a MWC may spend more time in a potentially risky humeral elevation range (30-60°) than the controls. The findings from this study do not support an effect of age, sex, pain, injury level, or years since injury on the humeral elevation workspace for adults with SCI who use a MWC. Future work should expand the understanding of loading of the upper extremity during daily life and characterize more in-depth information about shoulder workspace and activities of daily living across injury levels and groups with and without pain and pathology.

## Data Availability

Data available upon request.

## Acknowledgements

This publication was made possible by funding from the National Institutes of Health (grant no. R01 HD84423-01), Mayo Clinic Robert D. and Patricia E. Kern Center for the Science of Health Care Delivery, and the National Center for Advancing Translational Sciences (UL1 TR002377).

**Appendix**

## Appendix A: Defining sensor-to-segment alignment matrices

When using inertial sensors to quantify the movement of body segments, determining the sensor-to-segment alignment is critical to enable accurate quantification of the orientation of the body segment rather than the orientation of the sensor. In other words, the orientation of a body segment’s anatomical reference frame with respect to the sensor reference frame must be determined so that estimates of the sensor orientation can be used to understand body segment orientation. In our study, we utilize functional alignment movements, in which participants complete some known movements or poses for which at least one anatomical axis can be estimated in an inertial measurement unit’s (IMU’s) body-fixed frame of reference.

Our functional alignment poses and movements are as follows:

FA1. Sitting/standing upright with arms at sides
FA2. Right arm t-pose (Figure 1 Posture 2)
FA3. Dynamic ab/adduction of the right shoulder, keeping right arm movement in frontal plane
FA4. Right arm flexion pose (Figure 1 Posture 3)
FA5. Dynamic flexion/extension of the right shoulder, keeping right arm movement in sagittal plane
FA6. Left arm t-pose (similar to Figure 1 Posture 2)
FA7. Dynamic ab/adduction of the left shoulder, keeping left arm movement in frontal plane
FA8. Left arm flexion pose (similar Figure 1 Posture 3)
FA9. Dynamic flexion/extension of the left shoulder, keeping left arm movement in sagittal plane
FA10. Dynamic trunk flexion/extension, keeping movement of thorax in sagittal plane
FA11. Sitting/standing upright with arms at sides
FA12. Simulated wheelchair propulsion or arm swing

Using data (IMU measured accelerations and angular velocities) from the above poses and movements, we construct sensor-to-segment alignment matrices for the thorax, right arm, and left arm as described below.

For the thorax, we first use the average acceleration due to gravity measured by the torso-mounted IMU during FA1 and FA11 to define a body segment fixed z-axis (superior-inferior axis, 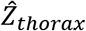) for the thorax:

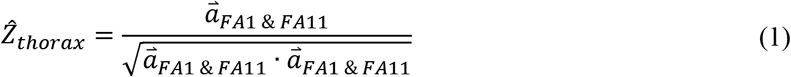

This establishes the thorax posture for which the thorax deviation angle is equal to zero. The average axis of rotation for the thorax (torso-mounted IMU) measured during FA10 is used to define the thorax x-axis (medial-lateral axis) for the thorax. To determine the average axis of rotation of the thorax segment (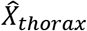), a principal components analysis is performed on the measured segment angular velocity measured during FA10. The first principal component is the unit vector defining the average axis of rotation.

We define an anterior-posterior axis (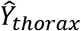) for the thorax as the unit vector orthogonal to the superior-inferior axis and medial-lateral axis:

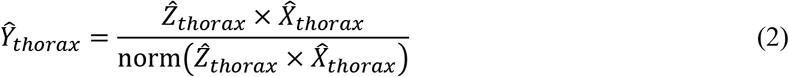

Finally, we ensure that the medial-lateral axis is orthogonal to the anterior-posterior and superior-inferior axes:

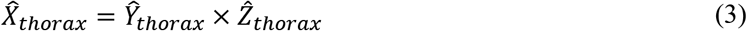

The resulting unit vectors (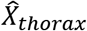, 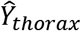, 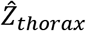) define a body-segment fixed frame aligned with estimated anatomical axes. The direction cosine matrix that defines the transformation of measurements made in the sensor-fixed frame to those in a thorax-fixed frame is defined by Equation (4), where each row of the matrix contains the components of the segment-fixed axes.

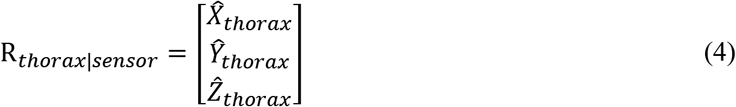

For the left and right arm, we aim to establish an axis that represents the long-axis of the humerus (defined here as the z-axis). Starting with the right arm, we first get one estimate of the z-axis of the upper arm by using acceleration measured during FA2 and angular velocity measured during FA3.

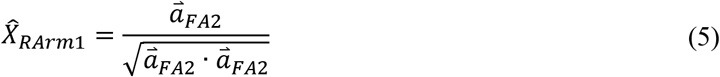

A principal components analysis is performed on the measured segment angular velocity measured during FA3; the first principal component is the unit vector defining the average axis of rotation during FA3 (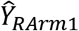). The first estimate of the z-axis of the upper arm is calculated:

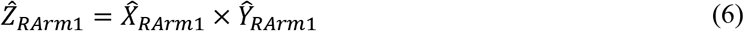

We choose 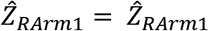 or 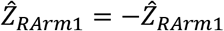 such that 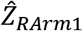 points superiorly (determined by examining 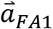 for the right arm). We get a second estimate of the z-axis of the upper arm by using acceleration measured during FA4 and angular velocity measured during FA5.

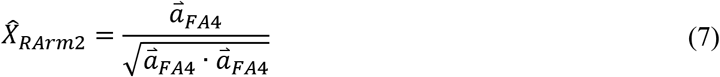

A principal components analysis is performed on the measured segment angular velocity measured during FA5; the first principal component is the unit vector defining the average axis of rotation during FA5 (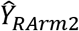). The second estimate of the z-axis of the upper arm is calculated:

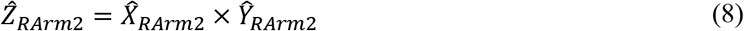

We choose 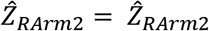 or 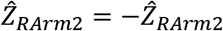 such that 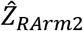 points superiorly (determined by examining 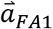 for the right arm).

We define the right arm z-axis using the two estimates:

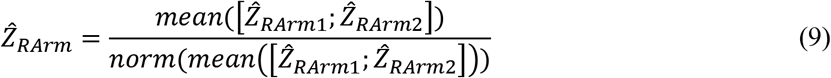

We construct a right arm x-axis:

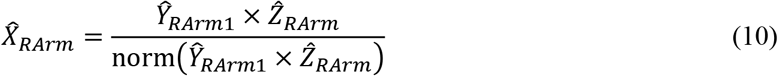

We choose 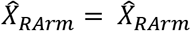 or 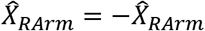 such that 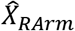 points up during FA2 (determined by examining 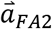 for the right arm).

Finally, we construct a right arm y-axis:

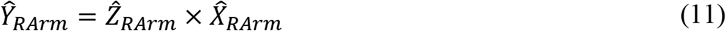

The resulting unit vectors (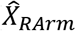, 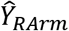, 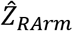) define a body-segment fixed frame aligned with estimated anatomical axes for the right arm. The direction cosine matrix that defines the transformation of measurements made in the sensor-fixed frame to those in a right arm-fixed frame is defined by Equation (12), where each row of the matrix contains the components of the segment-fixed axes.

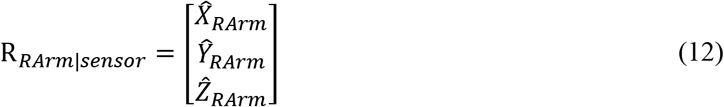

For the left arm we first get one estimate of the z-axis of the upper arm by using acceleration measured during FA6 and angular velocity measured during FA7.

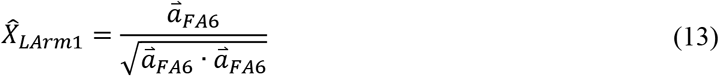

A principal components analysis is performed on the measured segment angular velocity measured during FA7; the first principal component is the unit vector defining the average axis of rotation during FA7 (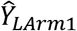). The first estimate of the z-axis of the upper arm is calculated:

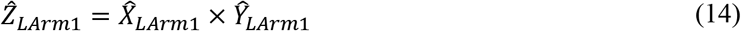

We choose 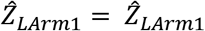 or 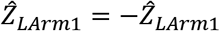 such that 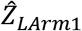 points superiorly (determined by examining 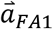 for the left arm). We get a second estimate of the z-axis of the upper arm by using acceleration measured during FA8 and angular velocity measured during FA9.

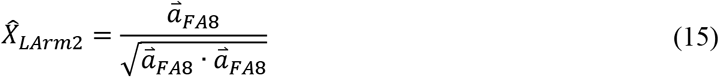

A principal components analysis is performed on the measured segment angular velocity measured during FA9; the first principal component is the unit vector defining the average axis of rotation during FA9 (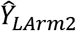). The second estimate of the z-axis of the upper arm is calculated:

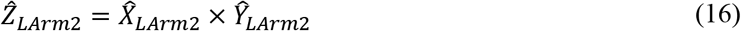

We choose 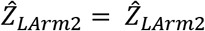 or 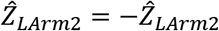 such that 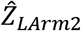 points superiorly (determined by examining 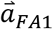 for the left arm).

We define the left arm z-axis using the two estimates:

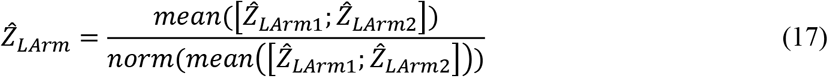

We construct a left arm x-axis:

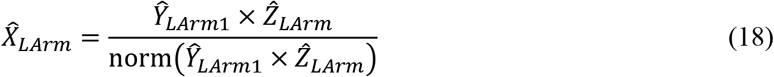

We choose 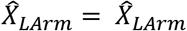 or 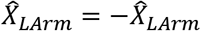 such that 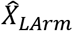 points down during FA6 (determined by examining 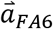 for the left arm).

Finally, we construct a left arm y-axis:

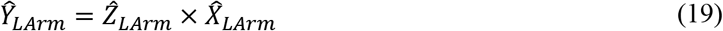

The resulting unit vectors (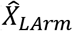, 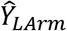, 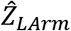) define a body-segment fixed frame aligned with estimated anatomical axes for the left arm. The direction cosine matrix that defines the transformation of measurements made in the sensor-fixed frame to those in a left arm-fixed frame is defined by Equation (19), where each row of the matrix contains the components of the segment-fixed axes.

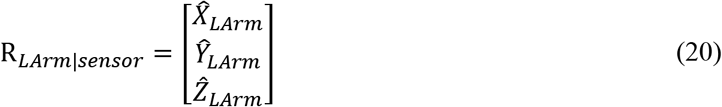

## Appendix B: Calculating humeral elevation thorax deviation angles

The humeral elevation angle and thorax deviation angle are defined as the angle between the segment z-axis and vertical and are calculated by first calculating the direction cosine matrix describing the orientation of the body segment in the inertial/world reference frame:

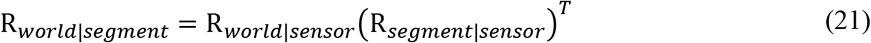

Where *R_world|sensor_* is the direction cosine matrix describing the rotation from the sensor reference frame to the inertial/world reference frame and *R_sensor|segment_* is the sensor-to-segment alignment from Equation (4), (12), or (20). Humeral elevation angles and thorax deviation angles are then calculated from Equation (22).

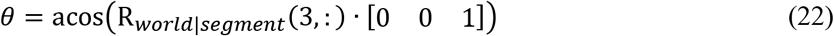

Where R*_world|segment_* refers to the last row of R*_world|segment_*.

## Appendix C: The distribution of the percentage of time the SCI and control cohort spent in 10° bins for both the dominant and non-dominant arms.

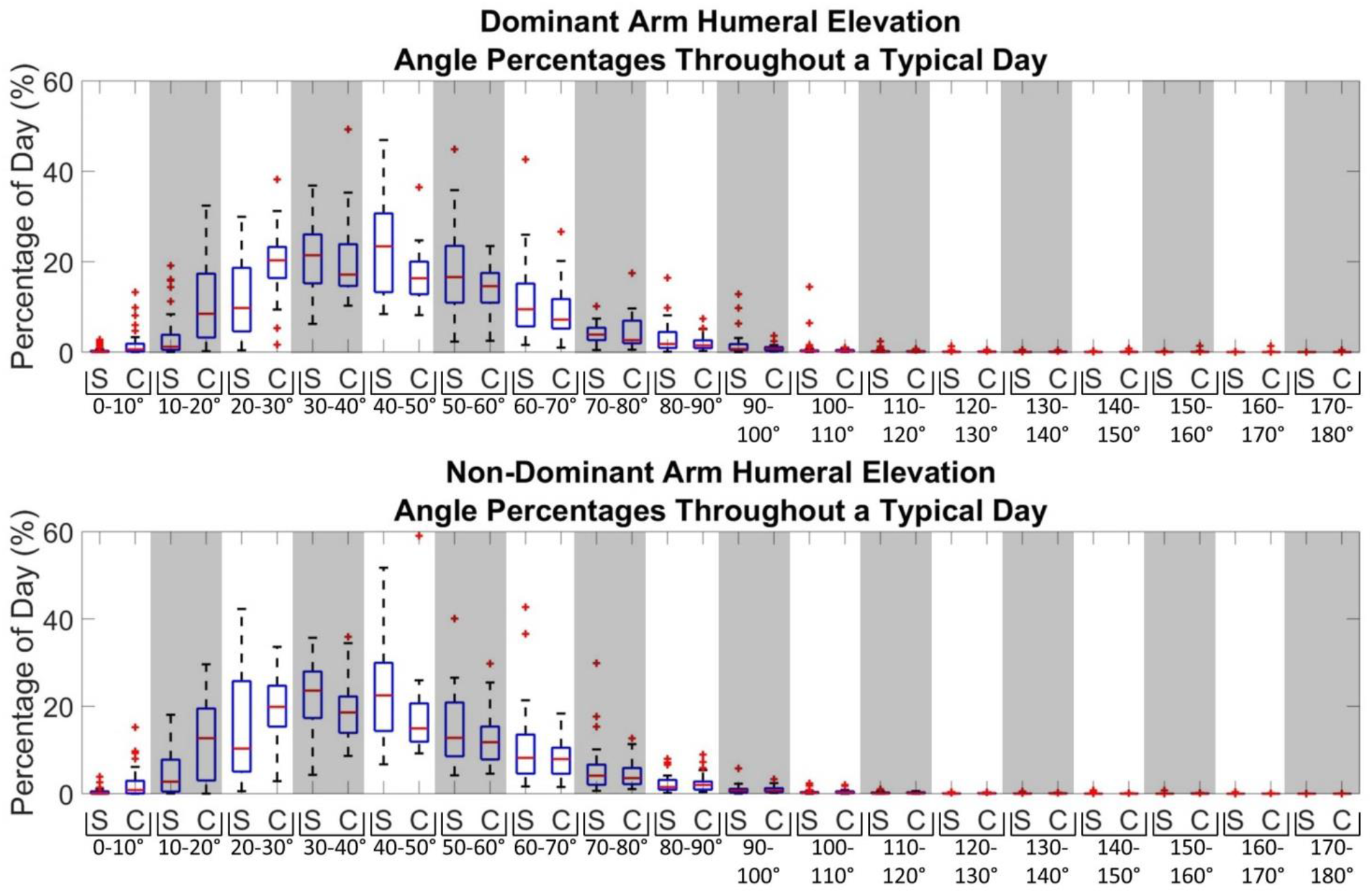

## References

1. Cooper RA, Boninger ML, Robertson RN. Repetitive Strain Injury Among Manual Wheelchair Users. Team Rehab Report. 1998;9(2):35–8.

2. Curtis KA, Drysdale GA, Lanza RD, Kolber M, Vitolo RS, West R. Shoulder pain in wheelchair users with tetraplegia and paraplegia. Archives of physical medicine and rehabilitation. 1999;80(4):453–7.

3. Alm M, Saraste H, Norrbrink C. Shoulder pain in persons with thoracic spinal cord injury: prevalence and characteristics. Journal of rehabilitation medicine. 2008;40(4):277–83.

4. Samuelsson K, Tropp H, Gerdle B. Shoulder pain and its consequences in paraplegic spinal cord-injured, wheelchair users. Spinal cord. 2004;42(1):41.

5. Dalyan M, Cardenas D, Gerard B. Upper extremity pain after spinal cord injury. Spinal cord. 1999;37(3): 191–5.

6. Divanoglou A, Augutis M, Sveinsson T, Hultling C, Levi R. Self-reported health problems and prioritized goals in community-dwelling individuals with spinal cord injury in Sweden. Journal of rehabilitation medicine. 2018;50(10):872–8.

7. Dyson-Hudson TA, Kirshblum SC. Shoulder pain in chronic spinal cord injury, part 1: epidemiology, etiology, and pathomechanics. Taylor & Francis; 2004.

8. Greving K, Dorrestijn O, Winters J, Groenhof F, Van der Meer K, Stevens M, et al. Incidence, prevalence, and consultation rates of shoulder complaints in general practice. Scandinavian journal of rheumatology. 2012;41(2):150–5.

9. Sie IH, Waters RL, Adkins RH, Gellman H. Upper extremity pain in the postrehabilitation spinal cord injured patient. Archives of physical medicine and rehabilitation. 1992;73(1):44–8.

10. Giner-Pascual M, Alcanyis-Alberola M, González LM, Aguilar-Rodriguez M, Querol F. Shoulder pain in cases of spinal injury: influence of the position of the wheelchair seat. International Journal of Rehabilitation Research. 2011;34(4):282–9.

11. Akbar M, Balean G, Brunner M, Seyler TM, Bruckner T, Munzinger J, et al. Prevalence of rotator cuff tear in paraplegic patients compared with controls. JBJS. 2010;92(1):23–30.

12. Seitz AL, McClure PW, Finucane S, Boardman III ND, Michener LA. Mechanisms of rotator cuff tendinopathy: intrinsic, extrinsic, or both? Clinical biomechanics. 2011;26(1): 1–12.

13. Carpenter JE, Flanagan CL, Thomopoulos S, Yian EH, Soslowsky LJ. The effects of overuse combined with intrinsic or extrinsic alterations in an animal model of rotator cuff tendinosis. The American journal of sports medicine. 1998;26(6):801–7.

14. Braman JP, Zhao KD, Lawrence RL, Harrison AK, Ludewig PM. Shoulder impingement revisited: evolution of diagnostic understanding in orthopedic surgery and physical therapy. Medical & biological engineering & computing. 2014;52(3):211–9.

15. Lawrence RL, Schlangen DM, Schneider KA, Schoenecker J, Senger AL, Starr WC, et al. Effect of glenohumeral elevation on subacromial supraspinatus compression risk during simulated reaching. Journal of Orthopaedic Research. 2017;35(10):2329–37.

16. Lawrence RL, Sessions WC, Jensen MC, Staker JL, Eid A, Breighner R, et al. The effect of glenohumeral plane of elevation on supraspinatus subacromial proximity. Journal of Biomechanics. 2018;79:147–54.

17. Giphart JE, van der Meijden OA, Millett PJ. The effects of arm elevation on the 3-dimensional acromiohumeral distance: a biplane fluoroscopy study with normative data. Journal of shoulder and elbow surgery. 2012;21(11): 1593–600.

18. Koontz AM, Cooper RA, Boninger ML, Souza AL, Fay BT. Shoulder kinematics and kinetics during two speeds of wheelchair propulsion. Journal of rehabilitation research and development. 2002;39(6):635–50.

19. Morrow MM, Kaufman KR, An KN. Scapula kinematics and associated impingement risk in manual wheelchair users during propulsion and a weight relief lift. Clinical biomechanics. 2011;26(4):352–7.

20. Requejo P, Mulroy S, Haubert LL, Newsam C, Gronley J, Perry J. Evidence-based strategies to preserve shoulder function in manual wheelchair users with spinal cord injury. Topics in Spinal Cord Injury Rehabilitation. 2008;13(4):86–119.

21. El-Gohary M, McNames J. Shoulder and elbow joint angle tracking with inertial sensors. IEEE Transactions on Biomedical Engineering. 2012;59(9):2635–41.

22. Bouvier B, Duprey S, Claudon L, Dumas R, Savescu A. Upper Limb Kinematics Using Inertial and Magnetic Sensors: Comparison of Sensor-to-Segment Calibrations. Sensors (Basel). 2015; 15(8): 18813–33.

23. Cutti AG, Giovanardi A, Rocchi L, Davalli A, Sacchetti R. Ambulatory measurement of shoulder and elbow kinematics through inertial and magnetic sensors. Med Biol Eng Comput. 2008;46(2): 169–78.

24. Kirking B, El-Gohary M, Kwon Y. The feasibility of shoulder motion tracking during activities of daily living using inertial measurement units. Gait & posture. 2016;49:47–53.

25. Chapman RM, Torchia MT, Bell J-E, Van Citters DW. Continuously monitoring shoulder motion after total shoulder arthroplasty: maximum elevation and time spent above 90° of elevation are critical metrics to monitor. Journal of shoulder and elbow surgery. 2019.

26. Langohr GDG, Haverstock JP, Johnson JA, Athwal GS. Comparing daily shoulder motion and frequency after anatomic and reverse shoulder arthroplasty. Journal of shoulder and elbow surgery. 2018;27(2):325–32.

27. Hudak PL, Amadio PC, Bombardier C, Beaton D, Cole D, Davis A, et al. Development of an upper extremity outcome measure: the DASH (disabilities of the arm, shoulder, and head). American journal of industrial medicine. 1996;29(6):602–8.

28. Beaton DE, Katz JN, Fossel AH, Wright JG, Tarasuk V, Bombardier C. Measuring the wole or the parts?: Validity, reliability, and responsiveness of the disabilities of the arm, shoulder and hand outcome measure in different regions of the upper extremity. Journal of Hand Therapy. 2001;14(2): 128–42.

29. Curtis K, Roach K, Applegate EB, Amar T, Benbow C, Genecco T, et al. Development of the wheelchair user’s shoulder pain index (WUSPI). Spinal Cord. 1995;33(5):290.

30. Curtis K, Roach K, Applegate E, Amar T, Benbow C, Genecco T, et al. Reliability and validity of the wheelchair user’s shoulder pain index (WUSPI). Spinal Cord. 1995;33(10):595.

31. Coley B, Jolles BM, Farron A, Aminian K. Arm position during daily activity. Gait & Posture. 2008;28(4):581–7.

32. Amasay T, Zodrow K, Kincl L, Hess J, Karduna A. Validation of tri-axial accelerometer for the calculation of elevation angles. International Journal of Industrial Ergonomics. 2009;39(5):783–9.

33. Amasay T, Latteri M, Karduna AR. In vivo measurement of humeral elevation angles and exposure using a triaxial accelerometer. Human factors. 2010;52(6):616–26.

34. Savage PG. Strapdown inertial navigation integration algorithm design part 1: Attitude algorithms. Journal of guidance, control, and dynamics. 1998;21(1): 19–28.

35. Sabatini AM. Quaternion-based extended Kalman filter for determining orientation by inertial and magnetic sensing. IEEE transactions on Biomedical Engineering. 2006;53(7):1346–56.

36. Madgwick S. An efficient orientation filter for inertial and inertial/magnetic sensor arrays. Report x-io and University of Bristol (UK). 2010;25:113–8.

37. McGinnis RS, Cain SM, Tao S, Whiteside D, Goulet GC, Gardner EC, et al. Accuracy of femur angles estimated by IMUs during clinical procedures used to diagnose femoroacetabular impingement. IEEE Transactions on Biomedical Engineering. 2015;62(6): 1503–13.

38. McGinnis RS, Cain SM, Davidson SP, Vitali RV, McLean SG, Perkins N, editors. Validation of complementary filter based IMU data fusion for tracking torso angle and rifle orientation. ASME International Mechanical Engineering Congress and Exposition; 2014: American Society of Mechanical Engineers.

39. Coley B, Jolles BM, Farron A, Aminian K. Detection of the movement of the humerus during daily activity. Medical & biological engineering & computing. 2009;47(5):467–74.

40. Granzow RF, Schall Jr MC, Smidt MF, Chen H, Fethke NB, Huangfu R. Characterizing exposure to physical risk factors among reforestation hand planters in the Southeastern United States. Applied Ergonomics. 2018;66:1–8.

41. Schall Jr MC, Fethke NB, Chen H. Working postures and physical activity among registered nurses. Applied ergonomics. 2016;54:243–50.

42. Chapman RM, Torchia MT, Bell J-E, Van Citters DW. Assessing shoulder biomechanics of healthy elderly individuals during activities of daily living using inertial measurement units: high maximum elevation is achievable but rarely used. Journal of biomechanical engineering. 2019; 141(4):041001.

43. An KN, Browne A, Korinek S, Tanaka S, Morrey B. Three-dimensional kinematics of glenohumeral elevation. Journal of Orthopaedic Research. 1991;9(1): 143–9.

44. Seel T, Schauer T, Raisch J, editors. Joint axis and position estimation from inertial measurement data by exploiting kinematic constraints. 2012 IEEE International Conference on Control Applications; 2012: IEEE.

45. Vitali R, Cain S, McGinnis R, Zaferiou A, Ojeda L, Davidson S, et al. Method for estimating three-dimensional knee rotations using two inertial measurement units: Validation with a coordinate measurement machine. Sensors (Basel). 2017;17(9):1970.

46. Kessel L, Watson M. The painful arc syndrome. Clinical classification as a guide to management. The Journal of bone and joint surgery British volume. 1977;59(2): 166–72.

47. McAtamney L, Corlett EN. RULA: a survey method for the investigation of work-related upper limb disorders. Applied ergonomics. 1993;24(2):91–9.

48. Lugade V, Fortune E, Morrow M, Kaufman K. Validity of using tri-axial accelerometers to measure human movement - Part I: Posture and movement detection. Med Eng Phys. 2014;36(2): 169–76.

49. Goodwin BM, Fortune E, Van Straaten MG, Morrow MM. Outcome Measures of Free-Living Activity in Spinal Cord Injury Rehabilitation. Current Physical Medicine and Rehabilitation Reports. 2019:1–6.

50. Sonenblum SE, Sprigle S, Lopez RA. Manual wheelchair use: bouts of mobility in everyday life. Rehabilitation research and practice. 2012;2012.

51. Tolerico ML, Ding D, Cooper RA, Spaeth DM. Assessing mobility characteristics and activity levels of manual wheelchair users. Journal of rehabilitation research and development. 2007;44(4):561.

52. Collinger JL, Boninger ML, Koontz AM, Price R, Sisto SA, Tolerico ML, et al. Shoulder biomechanics during the push phase of wheelchair propulsion: a multisite study of persons with paraplegia. Archives of physical medicine and rehabilitation. 2008;89(4):667–76.

53. Rao SS, Bontrager EL, Gronley JK, Newsam CJ, Perry J. Three-dimensional kinematics of wheelchair propulsion. IEEE Transactions on Rehabilitation Engineering. 1996;4(3): 152–60.

54. Collins SH, Adamczyk PG, Kuo AD. Dynamic arm swinging in human walking. Proceedings of the Royal Society B: Biological Sciences. 2009;276(1673):3679–88.

55. Fortune E, Cloud-Biebl BA, Madansingh SI, Ngufor CG, Van Straaten MG, Goodwin BM, et al. Estimation of manual wheelchair-based activities in the free-living environment using a neural network model with inertial body-worn sensors. Journal of electromyography and kinesiology. 2019.

56. Schneider S, Popp WL, Brogioli M, Albisser U, Demkó L, Debecker I, et al. Reliability of wearable-sensor-derived measures of physical activity in wheelchair-dependent spinal cord injured patients. Frontiers in neurology. 2018;9:1039.

